# The Impact of Cord Clamping on Haemodynamic Transition in Term Newborn Infants

**DOI:** 10.1101/2023.02.13.23285889

**Authors:** R. Chioma, D. Finn, D. Healy, V. Livingstone, J. Panaviene, E. M. Dempsey

## Abstract

**Background:** The transition from fetal to neonatal circulation is a complex physiological phenomenon, influenced by umbilical cord clamping and lung aeration, which triggers an increase in pulmonary blood flow and left ventricular preload. Although clinical evidence suggests that delayed cord clamping (DCC) prevents complications of haemodynamic instability, such as cerebrovascular injury, the cardiovascular consequences of DCC have not been investigated yet in humans.

**Methods:** Echocardiography was performed in 46 term vigorous infants before DCC, immediately after DCC, and at 5 minutes of life. Pulsed-wave Doppler-derived cardiac output and the pulmonary artery acceleration time indexed to the right ventricle ejection time, as a proxy of right ventricular afterload, were obtained. As markers of pre- and afterload fluctuations, the myocardial performance indexes and the velocities of the tricuspid and mitral valve annuli were determined with tissue Doppler imaging. Heart rate was derived from Doppler imaging and obtained throughout the assessments. Echocardiographic parameters were compared across the three timepoints using repeated measures ANOVA or Friedman’s test.

**Results:** DCC occurred at a median of 65 seconds (interquartile range 60-70). Left ventricular output increased throughout the first minutes of life (mean(SD): 222.4(32.5) mL/Kg/min before CC *vs*. 239.7(33.6) mL/Kg/min at 5 minutes, *P*=0.01), while right ventricular output dropped after CC (306.5(48.2) mL/Kg/min before CC *vs*. 272.8(55.5) mL/Kg/min immediately after CC, *P*=0.001). Right ventricular afterload rose after CC, decreasing in the following minutes. The tissue Doppler measurements showed that the loading conditions of both ventricles were transiently impaired by CC, recovering at 5 minutes. The heart rate progressively decreased after birth, following a linear trend temporarily disrupted by CC. Forward stepwise regression indicated that the variation in left ventricular output across the CC was directly correlated to the fluctuation of left ventricular preload over the same period (*P=*0.03).

**Conclusion:** This study unveils the cardiovascular consequences of DCC in term vigorous infants and offers insight into the haemodynamic transition from fetal to neonatal circulation in humans. Strategies that aim to enhance left ventricular preload before CC, such as initiating ventilation with an intact umbilical cord in apneic infants, may prevent complications of perinatal cardiovascular imbalance.

**CLINICAL PERSPECTIVE:** *What Is New?:*

- Delaying cord clamping after birth provides a smoother haemodynamic transition from fetal to neonatal circulation, although its cardiovascular impact has not been investigated yet in human infants.
- We showed that, despite a transient imbalance of biventricular loading conditions, the cardiac output remains stable throughout the early cardiovascular transition in term vigorous infants receiving delayed cord clamping, especially on the left side, which accounts for cerebral perfusion.
- The variation of left ventricular output across the cord clamping is independently influenced by the establishment of left ventricular preload.

*What Are The Clinical Implications?:*

- Our study reports the complex cardiovascular phenomena occurring after the birth of human infants, offering insight into the haemodynamic challenges of the transitioning heart.
- Strategies that aim to optimize the left ventricular preload during the early phases of life, such as establishing ventilation before clamping the umbilical cord, may prevent or mitigate complications of perinatal haemodynamic imbalance, such as cerebrovascular injury.

## INTRODUCTION

The transition from fetal to neonatal circulation is a complex and fascinating physiological phenomenon. The cardiorespiratory changes during the first seconds after birth are perhaps the most critical and dramatic encountered in humans. In utero, gas exchange occurs across the placenta, which acts as a low-resistance vessel receiving approximately half of the combined fetal ventricular output ^1^. The pulmonary circulation is characterized by high pulmonary vascular resistance (PVR) and low blood flow to the lung (approximately 10-20% of the combined cardiac output), and the majority of the right ventricular output (RVO) is diverted away from the lungs to other organs via the ductus arteriosus ^2,3^. Oxygenated blood arrives to the right atrium via the ductus venosus and passes through the foramen ovale to the left atrium ^4^. Thus, left ventricular (LV) preload is predominantly derived from the placental circulation.

The onset of breathing at birth, leading to lung aeration, is critical for unimpeded fetal to neonatal transition. This triggers a dramatic drop in PVR with a consequent immediate increase in pulmonary blood flow ^5^, crucial to allow alveolar gas exchange and the establishment of pulmonary venous return ^6^. Removal of the low-resistance placental circuit at the time of umbilical cord clamping (CC) ultimately determines the transition to the neonatal circulation. It results in an immediate increase in systemic vascular resistance (SVR) ^6^, and transiently reduces biventricular venous return following occlusion of the umbilical vein.

Time derived delayed CC (DCC) after birth in vigorous infants is the current international standard and is supported by a growing body of evidence ^7-9^. This intervention aims to facilitate placental blood transfusion to the infant, increasing hemoglobin levels at birth and improving iron stores in the first months of life ^10,11^, although some controversy exists around the timing and the extent of this transfusion ^12^. In preterm infants, DCC is associated with lower rates of intraventricular hemorrhage ^13^, presumably due to smoother cardiovascular adaptation and less haemodynamic disturbance ^14^. A recent animal study has provided further insight into the cardiovascular consequences of delaying the time of CC ^6^. In sedated lambs receiving positive pressure ventilation, haemodynamic benefits of clamping the cord after the establishment of pulmonary blood flow are seen. These include stabilization of carotid artery blood pressure and flow, prevention of bradycardia and sustained fall in RVO, and mitigation of increases in pulmonary artery pressure. Nevertheless, the evidence about post-natal haemodynamic transition is based on animal models, and data on the immediate cardiovascular consequences of DCC in human neonates is lacking. This study aimed to non-invasively monitor the transition from fetal to neonatal circulation and measure the cardiovascular changes associated with DCC in vigorous spontaneously breathing term infants born by elective cesarean section, through repeated targeted echocardiographic assessments immediately after birth.

## METHODS

This prospective observational study was conducted at Cork University Maternity Hospital (CUMH), Ireland between the 5^th^ of May and the 6^th^ of July 2022. CUMH is a tertiary university maternity hospital with more than 7000 deliveries annually, of which approximately 1100 are performed *via* elective cesarean section. The study was approved by the institutional research ethics board (Clinical Research Ethics Committee of the Cork Teaching Hospitals). Written parental consent was obtained from the parents on the antenatal wards before the elective cesarean section.

Mothers of singleton pregnancies with a gestational age above or equal to 37 weeks scheduled for elective cesarean section were eligible. Infants were excluded from the study in case of: *1)* contraindications to DCC, *2)* congenital heart defects, and *3)* chromosomal abnormalities.

Clinical data collected included the infants’ gestational age, birth weight, and gender. The timing of CC, Apgar scores at 1 and 5 minutes of life, body temperature after 10 minutes from birth, and the indication for elective cesarean delivery were also documented. In accordance with local guidelines, the umbilical cord was clamped at least 60 seconds after birth and cut by the obstetrician. Targeted echocardiograms were performed at three different time points by the same operator (RC): before CC (T1), immediately after CC (T2), and at approximately 5 minutes of life (T3). Immediately following delivery, the infant remained on the mother’s lower abdomen whilst DCC occurred, and for approximately one minute following the clamping of the cord. After completion of the second echocardiogram, they were moved to the resuscitation table. The clinical team, not involved in the research project, provided standard care throughout the study procedures. By local guidelines, skin-to-skin contact was performed starting approximately 10 minutes after birth.

### Echocardiography Measurements

At each time point, echocardiography was performed using a Vivid E95 ultrasound machine (GE Medical, Milwaukee, WJ) with a high frequency (8-12 MHz) neonatal probe. The measurements were performed according to current guidelines ^15,16^. The scans were stored in an online archiving system, and off-line analysis was carried out on the commercial software package supplied with the system. The pulsed wave Doppler measurements were obtained at the level of the aortic and pulmonary valve annuli (in the apical 5-chamber and the parasternal long axis views, respectively). All the Doppler recordings were obtained at an insonation angle <10° to the blood flow. Angle correction was not used, for feasibility reasons. Doppler indices were measured from the average of at least 3 consecutive cycles. The inner diameters (d) of pulmonary and aortic annuli were measured in the parasternal long axis view using two-dimensional echocardiography. The following features were determined: time velocity integral (TVI); stroke volume: (TVI × d^2^/4 × π)/birthweight; heart rate (HR); output per minute: stroke volume × HR. Pulmonary artery acceleration time (PAAT) was measured from the pulsed wave Doppler image of the blood flow through the pulmonary valve. This was indexed to the right ventricle ejection time (RVET) as follows: PAATi = PAAT/RVET. PAATi is a tricuspid regurgitation-independent and reliable measure of RV afterload ^17^. It is inversely proportional to the magnitude of the RV afterload. Tissue Doppler Imaging (TDI) velocities were obtained from the apical 4-chamber view. A pulsed wave Doppler sample gate of 2 mm was placed at the lateral tricuspid and lateral and septal mitral annuli. Peak myocardial systolic velocity (S’), and early (E’) and late (A’) diastolic velocities were measured from the average of at least 3 consecutive cycles. If the diastolic velocity waves were fused due to tachycardia, the single diastolic velocity was measured as A’. The isovolumic contraction time (interval between the end of the A’ to the onset of the S’), the isovolumic relaxation time (interval from the end of the S’ to the onset of the E’ of the following cardiac cycle), the ejection time (duration of S’), and the HR were measured on the TDI recordings. Myocardial performance index (MPI) was calculated using the formula: (isovolumic contraction time + isovolumic relaxation time)/ejection time. MPI is a measurement of global (systolic and diastolic) ventricular performance. It is directly proportional to the grade of ventricular dysfunction. All infants underwent a comprehensive echocardiogram at 24 hours of life to rule out congenital heart defects.

### Statistical Analysis

Categorical variables were described using frequency and percentage. Continuous variables were tested for normality with histogram illustrations and the Shapiro-Wilk test. Continuous variables were described using mean (standard deviation, SD) when normally distributed or median (interquartile range, IQR) otherwise. Normally distributed echocardiographic parameters were compared across the three time points using repeated-measures ANOVA. The Greenhouse-Geisser adjusted *P* value was reported when the assumption of sphericity was violated. Post-hoc pairwise comparisons were performed with Bonferroni adjustment. Skewed echocardiographic parameters were compared across the timepoints using the Friedman test followed by post-hoc pairwise Dunn-Bonferroni tests. The heart-rate trend across 15 timepoints (5 at each of T1, T2, and T3) was tested for linearity using repeated measurements ANOVA. The cumulative sum (CUSUM) test for structural breaks analysis was conducted on the progression of mean heart rate across the 15 timepoints. Forward stepwise multiple linear regression analyses were performed to investigate variables associated with variation (Δ) of LVO and RVO across the first two time points (T1-T2), expressed as the difference between the values before and after CC (ΔLVO and ΔRVO). The variables were selected *a priori*, based on physiological principles. Variables were inserted in the model if *P* was lower than 0.05. For the ΔRVO, the following variables were chosen: ΔS’, ΔA’, and ΔMPI measured at the lateral tricuspid annulus; and ΔPAATi. For the ΔLVO, the following variables were chosen: ΔS’, ΔA’, and ΔMPI measured at the lateral and septal mitral annuli. The TDI-derived E’ waves measured on tricuspid and mitral annuli were excluded from the analysis due to missing data, as the diastolic waves often were fused due to tachycardia. Statistical analysis was performed using IBM SPSS Statistics v 28 (Armonk, NY, USA: IBM Corp) and Prism 9 (GraphPad Software, San Diego, California). All tests were two-tailed, and a *P* value of <.05 was considered statistically significant.

We aimed to enroll a convenience sample of approximately 40 infants. A sample size of 40 infants would have a power of 94% to detect a medium effect (partial eta squared=0.06) in a repeated measures ANOVA with three timepoints, assuming a level of significance of 0.05, a 2-tailed test, correlations among repeated measures of 0.5 and a non-sphericity coefficient of 1.

## RESULTS

The clinical characteristics of infants included in the analysis are outlined in Table 1. Fifty infants born at term by elective cesarean section were recruited. Forty-six infants were included in the analysis. Three infants were excluded due to contraindication to DCC, and one infant due to unexpected chromosomal abnormality (see Figure 1). The median (IQR) gestation was 39.0 (38.7 – 39.2) weeks and the mean (SD) birth weight was 3498 (379) grams. Twenty-five (54%) infants were male. CC occurred at a median (IQR) of 65 (60 – 70) seconds, and Apgar scores were 9 (9 – 9) and 10 (10 – 10) at one and five minutes of life, respectively. All the infants were vigorous and actively breathing before the first echocardiogram, and none of them received CC before 60 seconds of life. The mean (SD) body temperature at 10 minutes after birth was 36.8 (0.17) degrees Celsius and no infant had a body temperature below 36.4 degrees C. Indications for elective cesarean section were: history of previous section (n=38, 83%), maternal request (n=4), maternal morbidities (n=3), and breech presentation (n=1).

**Table 1.** Characteristics of infants included in the analysis, n=46.

|  | Mean (SD), Median [IQR], or n (%) |
| --- | --- |
| Gestation, weeks | 39.0 [38.7 – 39.2] |
| Birth weight, g | 3498 (379) |
| Male, n | 25 (54%) |
| Timing of cord clamping, seconds | 65 [60-70] |
| 1 minute Apgar score | 9 [9-9] |
| 5 minutes Apgar score | 10 [10-10] |
| Body temperature at 10 minutes of life, degrees Celsius | 36.8 (0.17) |
| Indication for elective cesarean section |  |
| Previous section, n | 38 (83%) |
| Maternal request, n | 4 (9%) |
| Maternal morbidities, n | 3 (6%) |
| Breech presentation, n | 1 (2%) |

**Figure 1.**
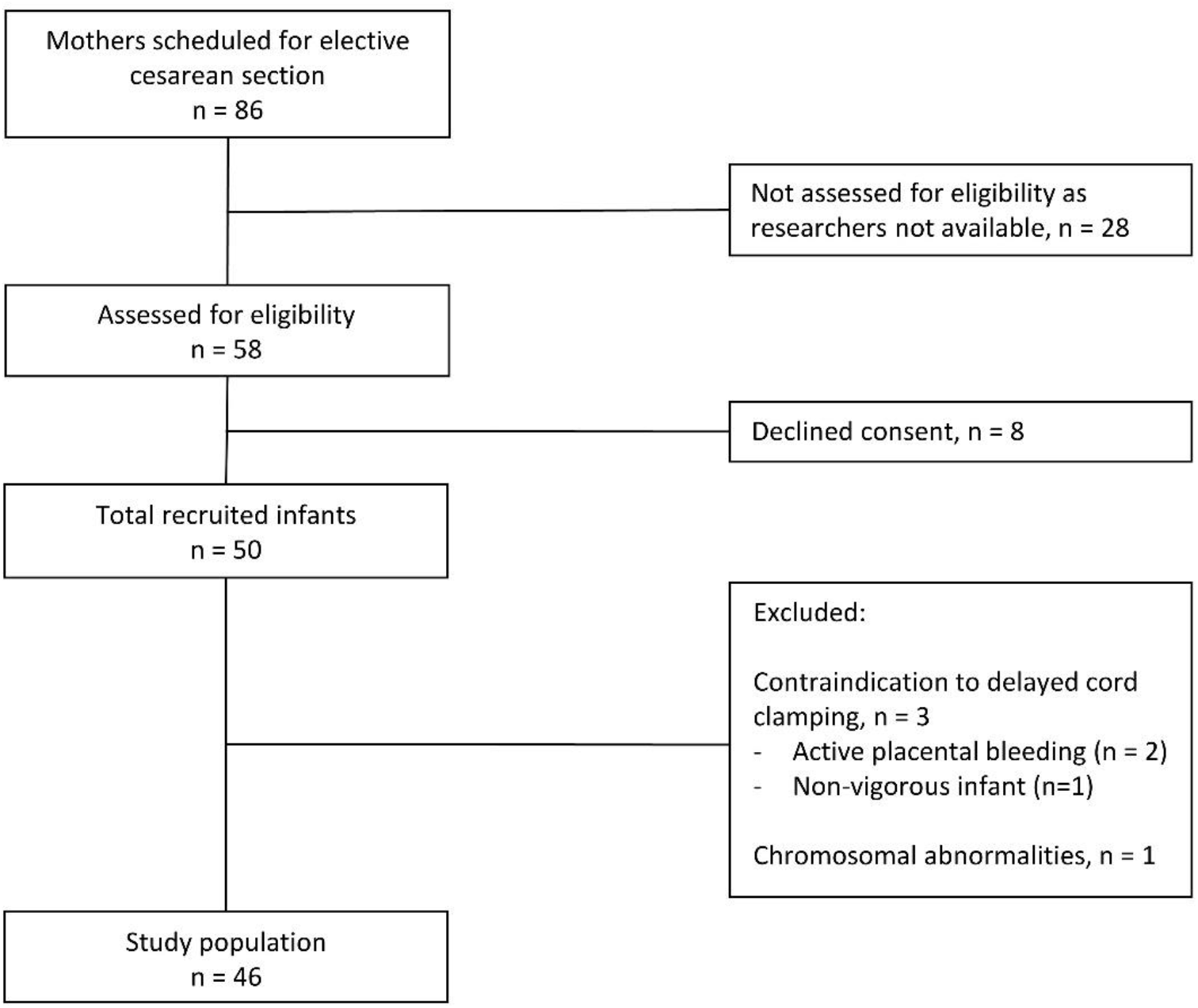
Study flow chart.

The serial echocardiography measurements are outlined in Table 2 and Figure 2. Heart rate progressively decreased after birth, following a linear trend (*P* <.001) over the singular measurements (5 at each of T1, T2, and T3), as shown in Figure 3. The CUSUM test revealed a structural break in this trend after CC (*P* = 0.013). The mean heart rate dropped throughout the three time points. RVO fell initially after CC and then stabilized at 5 minutes of life. The increase of RV stroke volume between T2 and T3 (mean(SD): 1.67 (0.30) mL/Kg and 1.76 (0.31) mL/Kg, respectively), although not statistically significant (pairwise comparison was not performed since the analysis of variance showed a *P* >.05), contributed to the stabilization of RVO, despite the decreasing heart rate. In contrast to this, the LVO significantly increased in the first 5 minutes of life (mean(SD): 239.7 (33.6) mL/Kg/min at T3 vs 222.4 (32.5) mL/Kg/min at T1, *P* =.01), due to the increase in the LV stroke volume, increasing significantly at each echo time point.

**Table 2.**
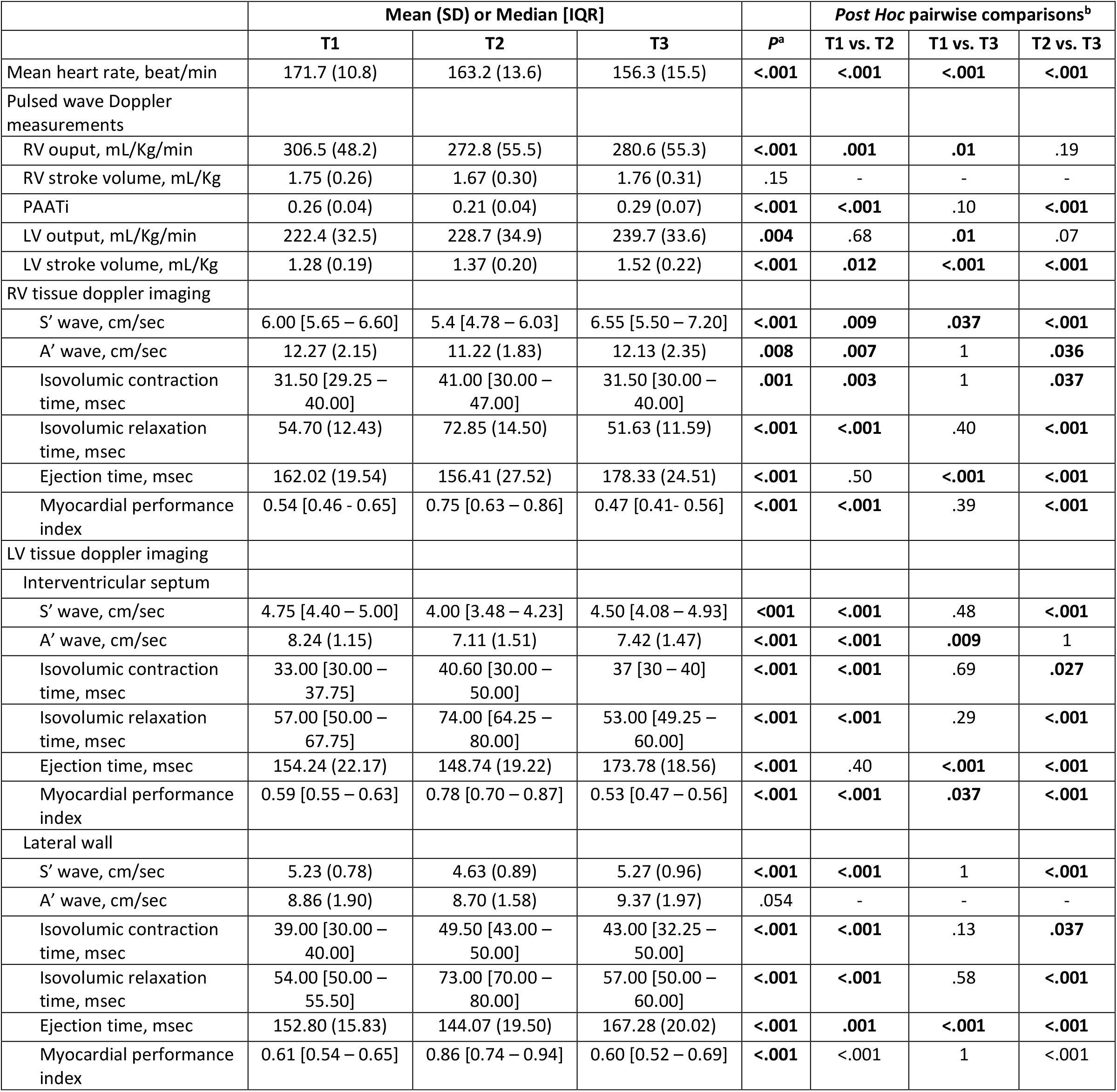
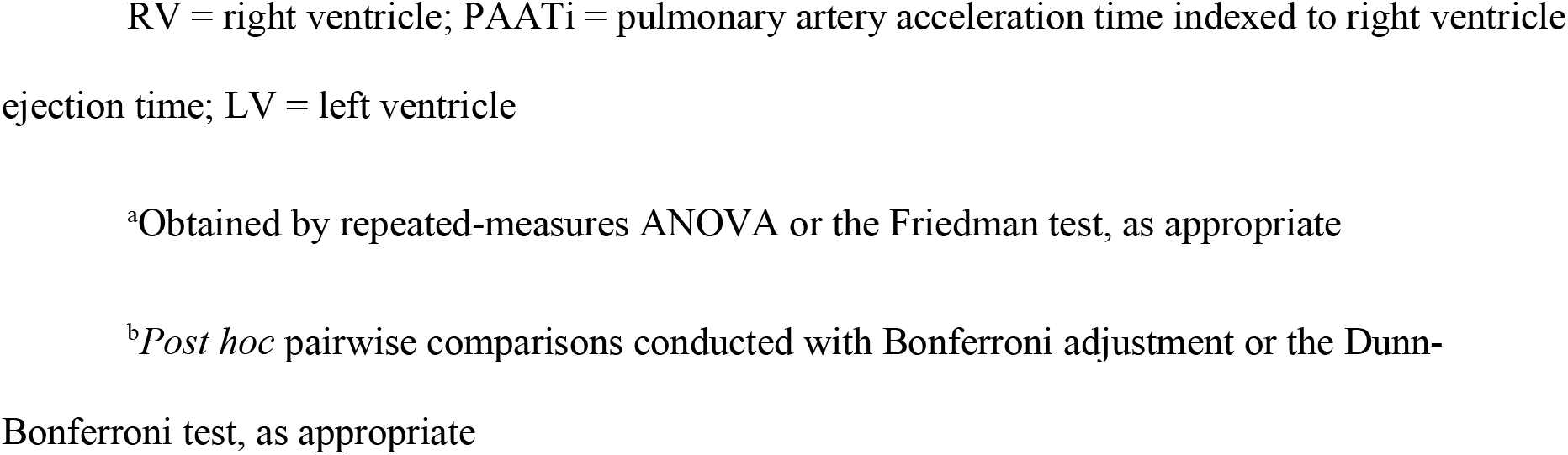
Echocardiographic measurements during three time points: before cord clamping (T1), after cord clamping (T2), and at 5 minutes of life (T3), n=46.

|  | Mean (SD) or Median [IQR] |  |  |  | Post Hoc pairwise comparisons<sup>b</sup> |  |  |
| --- | --- | --- | --- | --- | --- | --- | --- |
|  | T1 | T2 | T3 | P<sup>a</sup> | T1 vs. T2 | T1 vs. T3 | T2 vs. T3 |
| Mean heart rate, beat/min | 171.7 (10.8) | 163.2 (13.6) | 156.3 (15.5) | <.001 | <.001 | <.001 | <.001 |
| Pulsed wave Doppler measurements |  |  |  |  |  |  |  |
| RV output, mL/Kg/min | 306.5 (48.2) | 272.8 (55.5) | 280.6 (55.3) | <.001 | .001 | .01 | .19 |
| RV stroke volume, mL/Kg | 1.75 (0.26) | 1.67 (0.30) | 1.76 (0.31) | .15 | - | - | - |
| PAATi | 0.26 (0.04) | 0.21 (0.04) | 0.29 (0.07) | <.001 | <.001 | .10 | <.001 |
| LV output, mL/Kg/min | 222.4 (32.5) | 228.7 (34.9) | 239.7 (33.6) | .004 | .68 | .01 | .07 |
| LV stroke volume, mL/Kg | 1.28 (0.19) | 1.37 (0.20) | 1.52 (0.22) | <.001 | .012 | <.001 | <.001 |
| RV tissue doppler imaging |  |  |  |  |  |  |  |
| S' wave, cm/sec | 6.00 [5.65 – 6.60] | 5.4 [4.78 – 6.03] | 6.55 [5.50 – 7.20] | <.001 | .009 | .037 | <.001 |
| A' wave, cm/sec | 12.27 (2.15) | 11.22 (1.83) | 12.13 (2.35) | .008 | .007 | 1 | .036 |
| Isovolumic contraction time, msec | 31.50 [29.25 – 40.00] | 41.00 [30.00 – 47.00] | 31.50 [30.00 – 40.00] | .001 | .003 | 1 | .037 |
| Isovolumic relaxation time, msec | 54.70 (12.43) | 72.85 (14.50) | 51.63 (11.59) | <.001 | <.001 | .40 | <.001 |
| Ejection time, msec | 162.02 (19.54) | 156.41 (27.52) | 178.33 (24.51) | <.001 | .50 | <.001 | <.001 |
| Myocardial performance index | 0.54 [0.46 - 0.65] | 0.75 [0.63 – 0.86] | 0.47 [0.41- 0.56] | <.001 | <.001 | .39 | <.001 |
| LV tissue doppler imaging |  |  |  |  |  |  |  |
| Interventricular septum |  |  |  |  |  |  |  |
| S' wave, cm/sec | 4.75 [4.40 – 5.00] | 4.00 [3.48 – 4.23] | 4.50 [4.08 – 4.93] | <.001 | <.001 | .48 | <.001 |
| A' wave, cm/sec | 8.24 (1.15) | 7.11 (1.51) | 7.42 (1.47) | <.001 | <.001 | .009 | 1 |
| Isovolumic contraction time, msec | 33.00 [30.00 – 37.75] | 40.60 [30.00 – 50.00] | 37 [30 – 40] | <.001 | <.001 | .69 | .027 |
| Isovolumic relaxation time, msec | 57.00 [50.00 – 67.75] | 74.00 [64.25 – 80.00] | 53.00 [49.25 – 60.00] | <.001 | <.001 | .29 | <.001 |
| Ejection time, msec | 154.24 (22.17) | 148.74 (19.22) | 173.78 (18.56) | <.001 | .40 | <.001 | <.001 |
| Myocardial performance index | 0.59 [0.55 – 0.63] | 0.78 [0.70 – 0.87] | 0.53 [0.47 – 0.56] | <.001 | <.001 | .037 | <.001 |
| Lateral wall |  |  |  |  |  |  |  |
| S' wave, cm/sec | 5.23 (0.78) | 4.63 (0.89) | 5.27 (0.96) | <.001 | <.001 | 1 | <.001 |
| A' wave, cm/sec | 8.86 (1.90) | 8.70 (1.58) | 9.37 (1.97) | .054 | - | - | - |
| Isovolumic contraction time, msec | 39.00 [30.00 – 40.00] | 49.50 [43.00 – 50.00] | 43.00 [32.25 – 50.00] | <.001 | <.001 | .13 | .037 |
| Isovolumic relaxation time, msec | 54.00 [50.00 – 55.50] | 73.00 [70.00 – 80.00] | 57.00 [50.00 – 60.00] | <.001 | <.001 | .58 | <.001 |
| Ejection time, msec | 152.80 (15.83) | 144.07 (19.50) | 167.28 (20.02) | <.001 | .001 | <.001 | <.001 |
| Myocardial performance index | 0.61 [0.54 – 0.65] | 0.86 [0.74 – 0.94] | 0.60 [0.52 – 0.69] | <.001 | <.001 | 1 | <.001 |

RV = right ventricle; PAATi = pulmonary artery acceleration time indexed to right ventricle ejection time; LV = left ventricle
<sup>a</sup>Obtained by repeated-measures ANOVA or the Friedman test, as appropriate
<sup>b</sup>*Post hoc* pairwise comparisons conducted with Bonferroni adjustment or the Dunn-Bonferroni test, as appropriate

**Figure 2.**
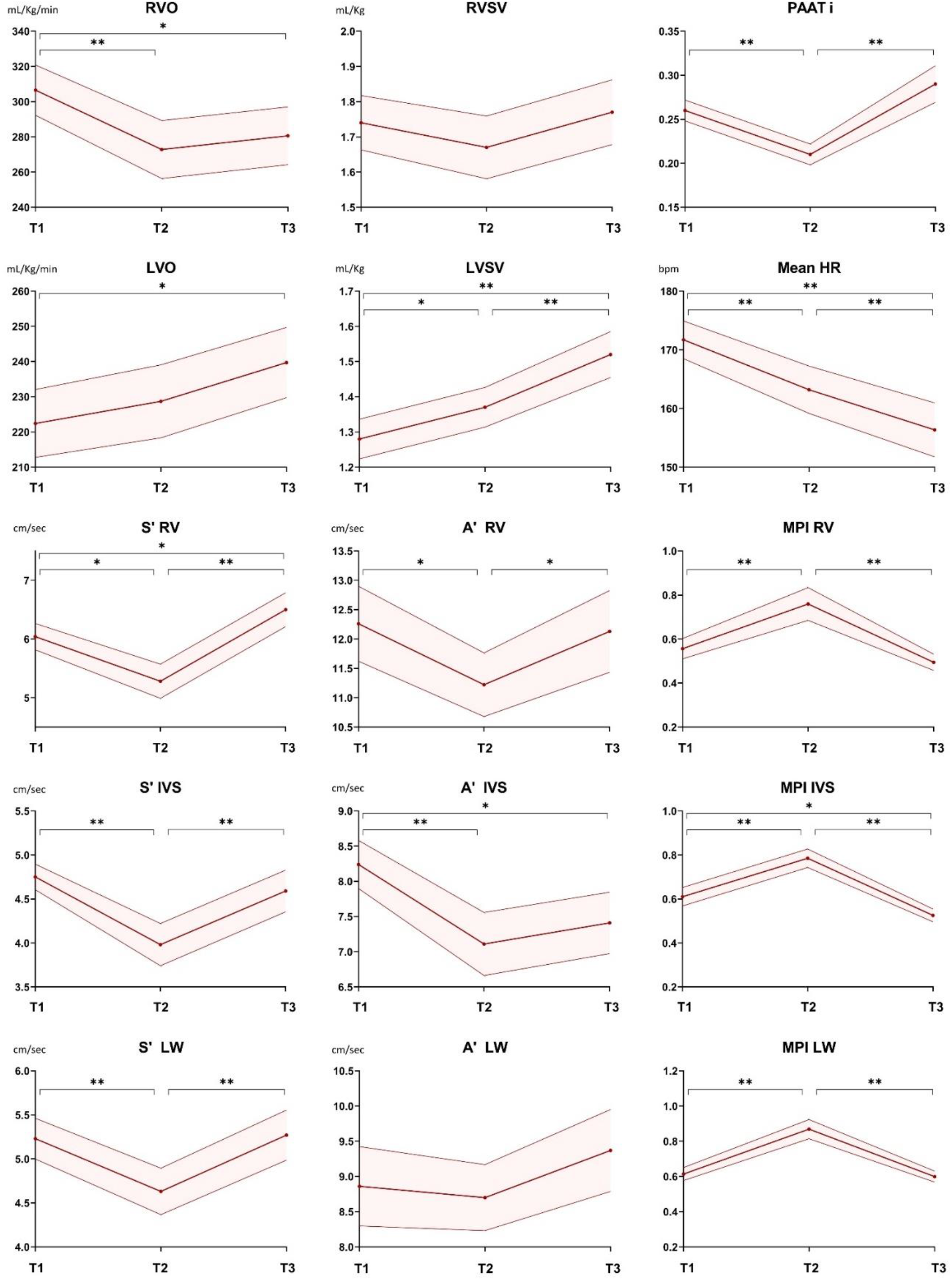
Non-invasive haemodynamic monitoring in transitioning term vigorous infants. Evolution of echocardiography measurements across the study time points: before cord clamping (T1), after cord clamping (T2), and at 5 minutes of life (T3). The dark red line joins the mean values across the time points, while the 95% confidence intervals are connected by the upper and lower line. RVO = right ventricular output; RVSV = right ventricular stroke volume; PAATi = pulmonary artery acceleration time indexed to right ventricle ejection time; LVO = left ventricular output; LVSV = left ventricular stroke volume; HR = heart rate; RV = right ventricle; MPI = myocardial performance index; IVS = interventricular septum; LW = lateral wall. \**P*<0.05, \*\**P*<0.001

**Figure 3.**
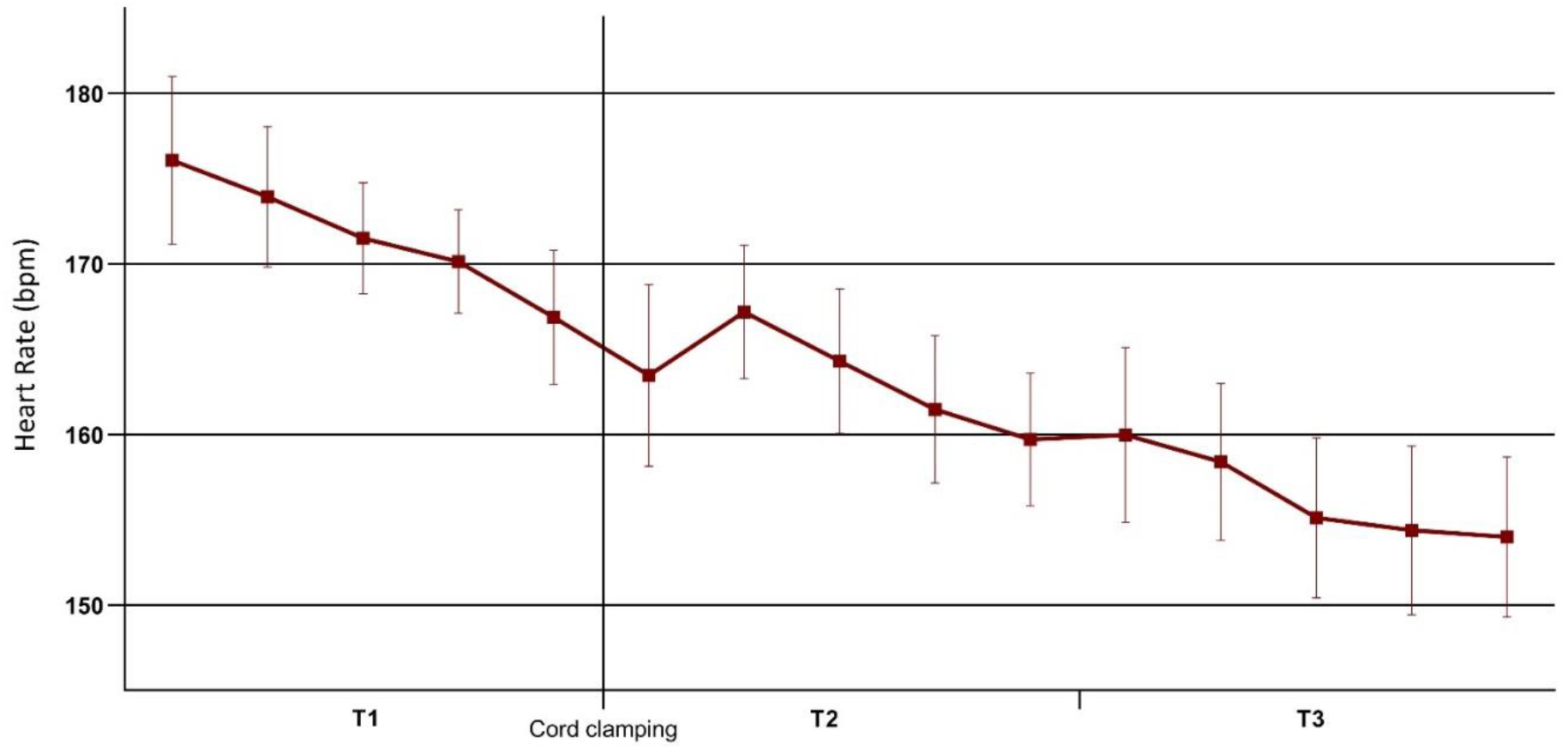
Heart rate trend after the birth of term vigorous infants and the influence of cord clamping. Detailed evolution of heart rate over the study time points: before cord clamping (T1), after cord clamping (T2), and at 5 minutes of life (T3). The dark red line joins the mean values across the time points, while the 95% confidence intervals were displayed with the error bars.

**Figure 4.**
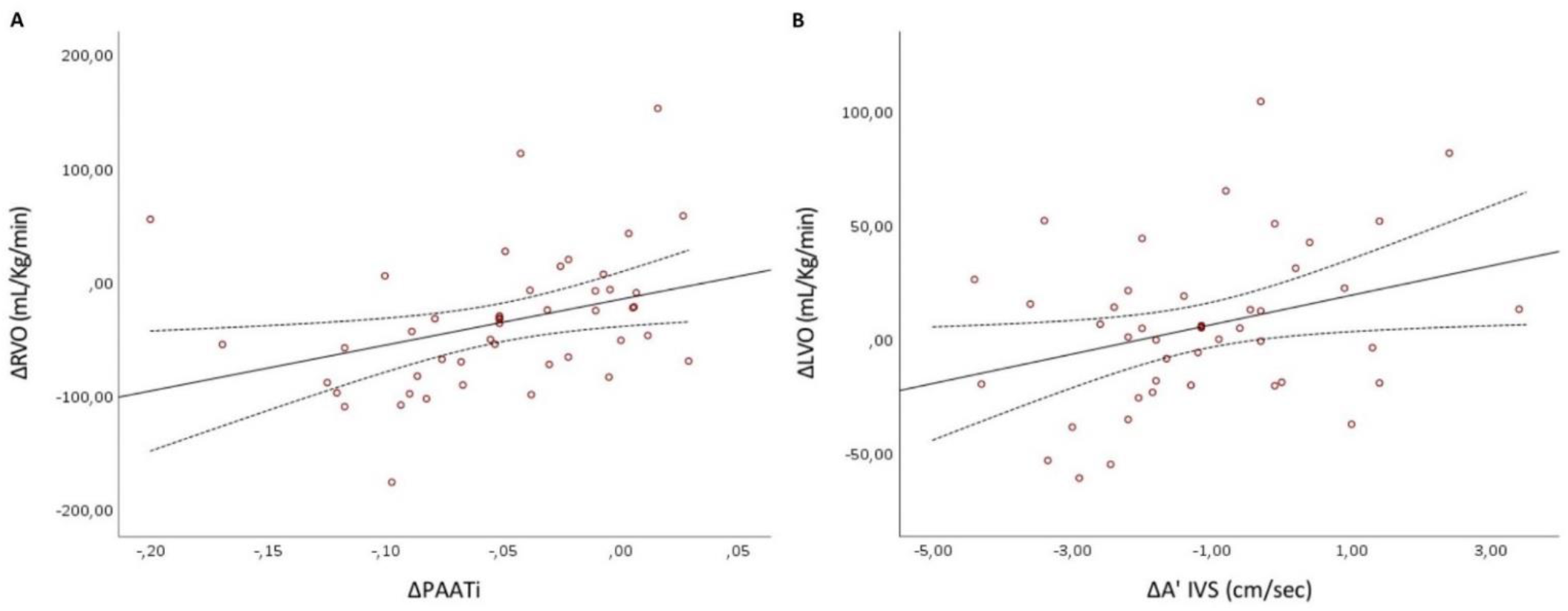
Association between variations of cardiac output across the cord clamping and variation of loading conditions over the same period. (A) Positive relationship between variation of right ventricular output (RVO) across the cord clamping and the variation of pulmonary artery acceleration time indexed to ejection time (PAATi) over the same time points. The stepwise regression model: ΔRVO = 13.29+402.51* ΔPAATi (B) Positive relationship between variation of left ventricular output (LVO) across the cord clamping and the variation of septal mitral annulus diastolic late velocity (A’ IVS) over the same time points. The stepwise regression model: ΔLVO = −13.57+6.48* ΔA’. Continuous lines represent the regression equation, while the dotted lines represent the 95% confidence intervals.

There was a reduction in PAATi from 0.26 (0.04) to 0.21 (0.04) immediately following CC (*P* <.001), which recovered to basal values at 5 minutes of life (0.29 (0.07)). This transient reduction represents an increase in RV afterload following CC. A significant decline occurred in the left and right ventricular TDI systolic velocities immediately after CC, improving at 5 minutes of life. A similar trend was followed by the TDI diastolic velocities of the tricuspid and septal mitral annuli, while the lateral mitral annulus was not affected by CC, not changing significantly throughout the early post-natal transition (*P* =.054). CC was associated with a transient and ubiquitous prolongation of the isovolumic contraction and relaxation times, which returned to baselines at T3. TDI-derived ejection times were not particularly influenced by CC and were significantly higher at 5 minutes of life compared to the previous time points. These variations resulted in a peak of left and right ventricular MPI after CC, dramatically dropping at T3.

For ΔRVO between T1 and T2 (T2-T1), the stepwise regression model was ΔRVO = 13.29+402.51* ΔPAATi, giving a Pearson’s correlation coefficient, r of 0.344, *P*=0.019. For ΔLVO, the stepwise regression model was ΔLVO = −13.57+6.48* ΔA’, giving r=0.315, *P*=0.033.

## DISCUSSION

In this study, we describe some key haemodynamic changes that occur immediately following birth in spontaneously breathing term infants delivered by cesarean section. To date, this is the first report investigating cardiovascular transition directly after birth and the haemodynamic consequences of CC in human neonates. We observed that: *1)* the HR progressively decreases after birth, following a linear trend transiently interrupted by CC; *2)* LVO and LV stroke volume increase throughout the first minutes of life, while RVO and RV stroke volume drop after CC; *3)* RV afterload rises after CC, significantly decreasing in the following minutes; *4)* TDI-derived myocardial velocities decline after CC, recovering at 5 minutes of life; *5)* the global ventricular performance temporarily falls after CC; *6)* the variation of LVO across CC is directly proportional to the variation of the septal mitral annulus A’ velocity (as a marker of LV preload) in the same timespan; *7)* the variation of RVO across CC is directly proportional to the variation of the RV afterload in the same time period.

Our findings of HR changes in the first minutes of life in human infants are somewhat novel, and perhaps the earliest recorded. Dawson et al reported HR trends retrospectively obtained by pulse oximetry monitoring in the first 10 minutes of life in three cohorts of well infants ^18^. They showed that, after reaching a peak at approximately 3-6 minutes of life, the HR slowly decreased in the following minutes. We observed consistently different HR values in the first minutes after birth, whereby the HR peak occurred in the first minute (at the first echocardiography assessment), and subsequently reduced thereafter. This could be explained by the different characteristics of the study populations, as Dawson et al. also included neonates born from vaginal delivery and did not stratify their results in accordance with factors that could influence HR in the very first minutes of life, such as the timing of CC and onset of respiration ^19,20^. Badurdeen et al recently developed HR percentiles between 1 and 10 minutes of life ^21^, detected by electrocardiography (ECG) from a cohort of infants born at ≥ 35 weeks’ gestation who had DCC and did not receive respiratory support after birth, stratifying their results based on the mode of delivery. They observed markedly higher HR values than previously reported by Dawson and colleagues, with a median of 170 bpm at 2 minutes after birth in cesarean deliveries, showing a subsequent decreasing trend. Our results support and extend these findings.

The accuracy of pulse oximetry is limited in the first minute of life, being influenced by motion artifacts, skin perfusion, and sensors’ instability on wet skin. This often leads to a delay that often exceeds 1-2 minutes after birth in obtaining a reliable HR signal ^22,23^. We determined HR from the direct visualization of the cardiac cycles on the Doppler ultrasound, as normally performed in fetal echocardiography ^3^. Cardiotocography is also based on Doppler ultrasound and is as accurate as direct fetal ECG in detecting HR ^24^. Thus, the mean HR in the first minute of life of spontaneously breathing infants born by cesarean section and receiving DCC seems consistently higher than previously assumed. HR values in the tachycardic range (> 180 bpm) appear to be a normal adaptation in the first minute of life, underlying the physiological stress caused by transitioning from fetal to neonatal circulation. Another novel finding was the impact of CC on HR, which caused a brief and transient HR increase. We speculate that this is an indirect sign of an acute reduction in biventricular preload following CC, triggering a temporary cardiac response to pseudo-hypovolemia.

As already observed in a previous report by van Vonderen et al. ^25^ who performed echocardiography assessing LV function at 2, 5, and 10 minutes of life in vigorous term neonates, LVO increased during the early haemodynamic transition, mainly determined by the significant and constant LV stroke volume increase. Our results are consistent with these findings, showing that LVO is not particularly affected by the cardiovascular consequences of CC. Considering the decreasing HR trend that we observed, the role of LV stroke volume appears crucial in the LV transition to neonatal circulation. In contrast, RVO was negatively affected by CC, dropping from a mean(SD) of 306.5 (48.2) mL/Kg/min before CC to 272.8 (55.5) mL/Kg/min immediately after CC. The main contributors were the reduced RV stroke volume coupled with a decreasing HR. The transient decrease in RVO following DCC is consistent with previously observed findings in preterm lamb studies ^6^.

The trend of PAATi is a novel finding, which confirms the close interplay between the RV and placental circulation in the early post-natal transition. PAATi has previously been validated as a reliable non-invasive marker of RV afterload in children and neonates ^17,26^. Interestingly, despite the expected fall in PVR due to lung expansion and aeration, we observed a transient increase in RV afterload immediately after CC, subsequently dropping at 5 minutes of life. In utero, ductal flow is right-to-left and facilitates the diversion of RVO to the lower body where a significant proportion is shunted across the placental circulation. In vigorous term infants, the direction of flow across the ductus arteriosus immediately after birth has been shown to resemble that of fetal circulation. Indeed, the ductus has about the same diameter as the descending aorta, being reported as (mean (SD)) 5.2 (1.3) mm at 2 minutes of life in a previous study on human neonates ^27^. As such, the increased RV afterload we have observed is most likely secondary to transductal propagation of the abrupt increase in SVR produced by clamping the umbilical arteries. To our knowledge, this is the first report showing the propagation of afterload oscillation produced by CC from the systemic to the pulmonary circulations. We also observed a significant drop in the RV afterload at T3, consistent with the progressive decrease in the PVR due to ongoing lung aeration and presumably enhanced pulmonary blood flow. The progressive increase in LV stroke volume during this time period supports enhanced pulmonary blood flow occurring.

TDI systolic and diastolic velocities acutely declined in both ventricles following CC, recovering at 5 minutes of life. TDI velocities are routinely employed as non-invasive proxy of longitudinal systolic and diastolic function. We selected these markers to assess the impact of CC on biventricular loading conditions, being particularly affected by acute changes in preload ^28-31^. Our data show that the preload of both ventricles is negatively affected by CC, even in infants with established ventilation. This means that RV and LV preload are still partially derived from the placental circulation at approximately 60 seconds of life in breathing babies. This finding is particularly relevant, as the interplay between CC and ventricular preload has been postulated but not yet demonstrated in previous works on the animal model ^6^. Interestingly, Drighil et al found that in young adults the lateral mitral annulus is more resistant to preload changes than either the lateral tricuspid annulus or the septal mitral annulus, being affected only by severe volume depletion ^32^. We observed that the lateral mitral TDI diastolic velocity was the only TDI index not influenced by CC, suggesting that its impact on LV preload was not substantial. At 5 minutes of life, the TDI velocities of both ventricles returned to pre-CC values, underlying the transient nature of this phenomenon.

MPI is a non-invasive Doppler marker of global ventricular performance that combines elements of both systolic and diastolic function. Increased RV and LV MPI have been identified as prognostic parameters characterizing patients with conditions such as pulmonary hypertension, cardiac failure, and myocardial infarction ^33-36^. Even though MPI has been reported to be relatively independent of heart rate and blood pressure ^37,38^, previous studies conducted on animal models observed that MPI is directly affected by acute changes in preload and afterload ^39,40^. Our data confirm these findings, as the documented variations of LV and RV MPI were caused by the previously discussed alterations in preload and afterload secondary to CC. Interestingly, at T2 we observed MPI considerably higher than reported normative values on the first day of life ^41^, highlighting the potentially significant physiological impact that CC has on the heart during adaptation. What is most reassuring is that on the follow-up scan at T3 we see a reversal of these changes and a return to baseline values.

Multiple linear regressions provided insights into the contribution of different haemodynamic factors playing a role in the transition from fetal to neonatal circulation. RVO variation across CC was independently associated with the variation of RV afterload. This result is not surprising, considering the aforementioned interplay between RV and placental circulation during the early haemodynamic transition and the afterload sensitivity of RV. LVO variation between T1 and T2 was independently associated with the variation of the septal mitral annulus A’ velocity. In other words, the less CC influences LV preload, the better the LVO trend. This finding indicates that the establishment of LV preload has a critical role in the post-natal transition of the left heart and highlights the importance of clamping the cord after ventilation initiation. This timing sequence enables pulmonary venous return to replace the umbilical venous return as the primary source of LV preload, guaranteeing the LV to smoothly assume its ex-utero role. This may be of particular clinical importance for delivery room stabilization of very preterm infants, who lack mature autoregulatory control of their cerebral circulation ^42,43^. Facilitating smooth LV transition, thereby minimizing swings in LVO, may reduce the risk of intraventricular hemorrhage for those infants.

The current recommendation in non-vigorous infants is to perform CC immediately after birth, prioritizing the initiation of resuscitation maneuvers ^9^. However, a recently published RCT suggests that this practice may not, in fact, be beneficial ^44^. It would be reasonable to assume that similar or more pronounced changes in cardiac performance occur with immediate CC compared to those described herein with DCC. As such, in circumstances where myocardial function may already be impaired, such as with birth asphyxia, the additional physiological stress to the myocardium caused by immediate CC may worsen rather than improve outcomes. In particular situations, performing resuscitation while the infant remains attached to the umbilical cord may facilitate smoother transition. Preliminary clinical trials demonstrate that initiating ventilation with an intact umbilical cord is feasible ^45,46^. This has also been demonstrated to improve haemodynamic stabilization in the setting of extreme prematurity ^47,48^. Considering the lower functional capacity of the immature heart to acutely cope with adverse loading conditions ^49^, these management strategies may be particularly relevant taken in the context of the previously discussed physiological changes associated with CC.

We acknowledge limitations of the present study. Given the restricted amount of time to perform these exceptionally early non-invasive measurements of cardiac function, we had to accurately target the echocardiography assessment. Hence, for feasibility reasons, we decided to exclude markers of blood shunting across the foramen ovale or the ductus arteriosus, and therefore, we could not directly document the interaction between systemic and pulmonary circulations or the interatrial shunting before and after CC. For the same reason, but also to limit the number of investigators operating in a sterile field, we could not include non-invasive measurements of cerebral oxygenation or systemic blood pressure, to monitor their trend during the early haemodynamic transition. For ethical reasons, considering the evidence supporting DCC, we could not design a comparative study to investigate the haemodynamic difference between immediate and delayed CC. To maintain homogeneous CC timing we did not include neonates born from vaginal delivery in the present study, who undergo CC typically at approximately 3 minutes of life in line with local policy drawn in accordance with the American College of Nurse-Midwives position statement ^8^. These aspects limit the generalizability of our results. Further studies are warranted to investigate the cardiovascular impact of CC in preterm and/or apneic infants and to assess the interaction between ventilation and CC in human neonates.

To conclude, our results underline the challenges of haemodynamic transition in well term infants, highlighting the importance of DCC. We showed that HR values in the first minutes after birth are higher than previously assumed, having a linear decreasing trend briefly disrupted by CC. Furthermore, CC produces a transient imbalance of biventricular loading conditions. RV afterload increased after CC, despite an expected drop in PVR, presumably caused by the transductal propagation of the increase in SVR. This phenomenon influenced RV transition from fetal to neonatal circulation, characterized by the drop in RVO due to interruption of placental shunting. The stabilization of LVO seems correlated to the establishment of LV preload, underlying the key role of lung aeration for the haemodynamic transition. Hence, we believe that the current recommendation in non-vigorous infants to perform CC immediately after birth warrants further consideration.

## Data Availability

The data that support the findings of this study are available upon request from the authors.

## Nonstandard Abbreviations and Acronyms

PVR: pulmonary vascular resistance
RVO: right ventricular output
LV: left ventricular
RV: right ventricular
SVR: systemic vascular resistance
DDC: delayed cord clamping
CC: cord clamping
TVI: time velocity interval
HR: heart rate
PAAT: pulmonary artery acceleration time
RVET: right ventricle ejection time
PAATi: pulmonary artery acceleration time indexed for right ventricle ejection time
TDI: tissue doppler imaging
MPI: myocardial performance index

## Acknowledgments

The authors were responsible for all content and editorial decisions, were involved at all stages of development, and approved the final version. Dr Chioma performed data collection and curation. Drs Chioma, Finn, Healy, and Prof Dempsey drafted the manuscript. Drs Chioma and Livingstone performed statistical analyses. All authors reviewed and edited the manuscript critically and approved the final version.

